# Prognostic Utility of Total Kidney Volume for Chronic Kidney Disease Risk Prediction: An Observational and Mendelian Randomization Study

**DOI:** 10.1101/2023.08.23.23294447

**Authors:** Jianhan Wu, Yifan Wang, Caitlyn Vlasschaert, Ricky Lali, James Feiner, Pukhraj Gaheer, Serena Yang, Nicolas Perrot, Michael Chong, Matthew B. Lanktree, Guillaume Paré

## Abstract

**Importance:** Low total kidney volume (TKV) is a risk factor for chronic kidney disease (CKD). However, evaluations of causal inference and prognostic utility beyond traditional biomarkers are lacking.

**Objective:** To investigate the observational and Mendelian randomization (MR) association of TKV with kidney and cardiovascular traits and assess improvement in CKD risk prediction when TKV is added to estimated glomerular filtration rate (eGFR) and urine albumin creatinine ratio (uACR).

**Design:** Case-control study based on UK Biobank and two-sample MR analysis.

**Setting:** 22 assessment centers, United Kingdom.

**Participants:** Individuals of European ancestry with kidney volume assessment derived from magnetic resonance imaging. Exclusion criteria include records of kidney transplantation, excision, congenital malformation, and cystic kidney disease.

**Exposures:** TKV, height adjusted (htTKV), and body surface area adjusted TKV (BSA-TKV).

**Main Outcomes:** Observational and bidirectional MR association estimates of TKV with CKD risk. Incident CKD stage G3 or worse prediction performance using likelihood ratio, C-statistic, calibration, and category-free net absolute reclassification index (NARI).

**Results:** Observational analysis included 34,595 individuals [median (IQR) age 64 (12) years, 17,835 (51.6%) females]. Adjusted for confounders and risk factors including eGFR and uACR, a 10 mL decrease in TKV was associated with 7% increase in the risk of incident CKD stage G3 or worse (HR 1.07, 95% CI 1.04 to 1.10, P < 0.001). Addition of prognostically significant BSA-TKV thresholds of 119 and 145 mL/m^2^ led to the greatest improvement in prediction performance beyond eGFR and uACR across likelihood ratio, discrimination (C-statistic 0.87, 95% CI 0.85 to 0.89, P = 0.017), calibration, and reclassification (NARI 228 per 1,000, P < 0.001). In MR, a 10 mL decrease in genetically predicted TKV was associated with 10% increase in CKD risk (OR 1.10, 95% CI 1.05 to 1.15, P < 0.001). Reciprocally, an increased risk of genetically predicted CKD by 2-fold was associated with an 8.75 mL reduction in TKV (95% CI -10.8 to -6.66, P < 0.001). There were no significant observational or MR associations of TKV with cardiovascular complications.

**Conclusions:** A bidirectional relationship exists between TKV and CKD. Addition of TKV thresholds to eGFR and uACR can improve CKD risk stratification.

## Introduction

The burden of chronic kidney disease (CKD) is large and growing, affecting more than 10% of the adult population resulting in significant morbidity and mortality.^1^ There is a need to improve risk stratification for CKD to identify those who require intensive intervention while re-assuring those at low risk of progression to kidney failure. Kidney imaging may improve upon estimated glomerular filtration rate (eGFR) and urinary albumin-to-creatinine ratio (uACR) for predicting incident kidney disease and complications.

Automated measurement of total kidney volume (TKV) can be obtained from magnetic resonance imaging (MRI).^2^ Height-adjusted total kidney volume (htTKV) has arisen as the most valuable prognostic biomarker in autosomal dominant polycystic kidney disease (ADPKD) as it correlates with cyst burden.^3^ In CKD, studies have reported a positive correlation between TKV and eGFR.^4–6^ Larger volumes are presumed to represent greater nephron endowment leading to increased glomerular filtration rate.^7,8^ Correspondingly, patients with advanced CKD show significant shrinkage of the kidneys as a result of atrophy.^9^ However, the causal directionality between TKV and CKD has not been tested beyond observational studies.

Here, we evaluated the association of TKV with kidney function measures and outcomes in 34,595 individuals, identifying prognostically significant thresholds to improve incident CKD risk prediction. We further analyzed cardiovascular complications which are the leading causes of mortality among those with CKD.^10^ Using two-sample Mendelian randomization (MR), we tested the genetic effect of TKV on kidney and cardiometabolic traits followed by reversing the exposure-outcome direction to assess their reciprocal effect on TKV. The results of MR analyses can infer causality between kidney volume and outcomes when key assumptions are met.^11^

## Methods

### Study Cohort

UK Biobank (UKB) is a multi-ethnic, prospective population cohort consisting of approximately 500,000 participants between the ages of 37-73 years.^12^ Individuals were recruited from the UK National Health Services between 2006 and 2010. Data pertaining to longitudinal health outcomes and cross-sectional sociodemographic, lifestyle, physical, and molecular traits were collected according to protocols described previously.^12^

Abdominal images were acquired for a subset of the UKB cohort using Siemens Aera 1.5T MRI scanner following the Dixon protocol.^13^ Participants were recruited on a voluntary basis but individuals with MRI contraindications were excluded. Kidney volumes were sourced from a previous study utilizing machine learning segmentation.^2^ Analysis was limited to individuals of European ancestry due to limited numbers in other ancestries. A total of 173 individuals with records of kidney transplantation, total or partial excision (OPCS Classification of Interventions and Procedures or OPCS4 M01 to M03), congenital malformations of the kidney (International Classification of Diseases, Tenth Revision, Clinical Modification or ICD10 Q60, Q63), and cystic disease (ICD10 Q61) or carriers of *PKD1* or *PKD2* loss of function variants were excluded from analysis, leaving a study cohort of 34,595 individuals.

### Outcomes and Exposures

All outcomes, exposures, and covariates are described in eTable 1 and 2. Summary of continuous variables are expressed in median and interquartile range (IQR). The primary outcome CKD stage G3+ was defined by any records of CKD above stage 3 (ICD10 N18.0, 18.3 to N18.5), dialysis (ICD10 Y60.2, Y84.1, Z49.0, Z49.1, Z49.2, Z99.2; OPCS4 X40 to X42), algorithmically defined end-stage kidney disease (UKB FID 42027), eGFR <60 mL/min/m^2^, or uACR >30 mg/mmol. Incidence were determined based on date of imaging assessment. TKV was height-adjusted (htTKV) or body surface area-adjusted (BSA-TKV) by dividing by height (m) or BSA (m^2^) based on the DuBois equation.

### Statistical Analysis

Incident disease outcomes were assessed using Cox proportional hazards models, censored by date of death or last follow-up. Prevalent disease outcomes were assessed using logistic regression and continuous traits were assessed using generalized linear models. Nonlinear associations were assessed using the multivariate fractional polynomial (MFP) method.^14^ Prognostically significant thresholds were identified based on a modified Mazumdar method.^15,16^ Models associating incident CKD stage G3+ with thresholds were iteratively compared to models without thresholds or the previous model of best fit using the likelihood ratio test. 1,000 iterations of bootstrap resampling was performed to assess threshold bias, standard deviation, and confidence interval. Thresholds were considered distinct if they fall outside of the 95% confidence intervals of other thresholds.

Multivariable models were adjusted for 1) age, sex, body size including body mass index (BMI) and BSA, first 10 genetic principal components of ancestry, image analysis protocol, image assessment center, 2) additionally smoking status, alcohol consumption, hemoglobin A1C (HbA1C), diabetes mellitus (DM), systolic blood pressure (SBP), and hypertension, and 3) additionally eGFR and uACR. Missing values were imputed using the multivariate imputation by chained equation predictive mean matching method.^17^

Improvement in prediction of incident CKD stage G3+ was assessed using the likelihood ratio test comparing nested models including eGFR and uACR with and without TKV. Discrimination was evaluated using C-statistic and significance of differences were assessed using the DeLong test. Calibration was assessed using the Hosmer-Lemeshow test which evaluates significance of differences between predicted and observed risk. Net absolute reclassification index (NARI) was calculated as the product of net reclassification index and event or non-event rate.^16^ Results were multiplied by 1,000 to obtain overall improvement per 1,000 individuals. Measurements were corrected for bias using Harrell’s bias correction with 500 iterations of bootstrap resampling.^18^

Genetically predicted TKV was calculated as the allelic sum of independent (r^2^ < 0.001) genetic variants associated beyond genome-wide significance level (P < 5 × 10^−8^) weighted by regression coefficients (eTable 3), based on a previous genome-wide association study (GWAS) by Liu et al.^2^ Association between genetically predicted TKV and traits were estimated using GWAS summary statistics using two-sample MR. A summary of all traits, assessment criteria, studies, ancestry, sample size, and UKB independence is available in eTable 4. Where available, independent cohorts without UKB overlapping samples were used as the main outcome. Validity of MR results rely on the assumptions of relevance, independence, and exclusion.^11^ The weighted-median method was used as the main method due to its consistent estimation in cases where up to 50% of the genetic variants are invalid instruments.^19^ Inverse-variance weighted was performed as secondary analysis. MR-Egger intercept test was performed to assess directional pleiotropy.^20^ Additionally, analyses were repeated with outlier variants, identified by the pleiotropy residual sum and outlier (MR-PRESSO) method^21^ and BMI or BSA-associated variants identified by NHGRI-EBI GWAS catalogue search, removed.

All analyses were performed in R (v 4.2.3). Statistical significance was considered if two-sided P-values are less than 0.05. P-values were corrected for multiple hypotheses testing using Bonferroni correction. This study followed Strengthening the Reporting of Observational Studies in Epidemiology (STROBE) guidelines for observational and MR analyses.

## Results

The study cohort consists of 34,595 individuals of European ancestry. 17,835 (51.6%) participants were female. Median (IQR) age at time of imaging is 64 (12) years. Median TKV is 266 mL (74.5; eFigure 1, eTable 5), median htTKV is 158 mL/m (37.6), and median BSA-TKV is 145 mL/m^2^ (29.2). There were 1,805 (5.2%) individuals with either prevalent or incident cases of CKD stage G3+ and 499 (1.4%) individuals with an acute kidney injury (AKI) event. Median eGFR_Cr_ and eGFR_CrCys_ were 98.7 (17.6) and 98.0 (15.8) mL/min/1.73m^2^ respectively.

Adjusted for age, sex, body size, first 10 genetic principal components, image analysis protocol version and assessment centers, a 10 mL reduction in TKV was significantly associated with 16% increase in risk of incident CKD stage G3+ (HR 1.16, 95% CI 1.13 to 1.19, P < 0.001) and 3% increase in risk of incident AKI (HR 1.03, 95% CI 1.01 to 1.06, P = 0.003) (Figure 1A). In parallel, a 10 mL reduction in TKV was significantly associated with 1.01 mL/min/1.73m^2^ reduction in eGFR_Cr_ (95% CI -1.04 to -0.99, P < 0.001) and 1.24 mL/min/1.73m^2^ reduction in eGFR_CrCys_ (95% CI -1.27 to -1.22, P < 0.001) (Figure 1B). We also observed a 0.01 mg/mmol reduction in uACR (95% CI -0.02 to 0.00, P = 0.03) per 10 mL reduction in TKV. In contrast, TKV was not significantly associated with incident cardiovascular outcomes including ischemic heart disease (HR 1.00, 95% CI 0.98 to 1.01, P = 0.89) or cerebrovascular disease (HR 1.00, 95% CI 0.99 to 1.02, P = 0.64).

**Figure 1:**
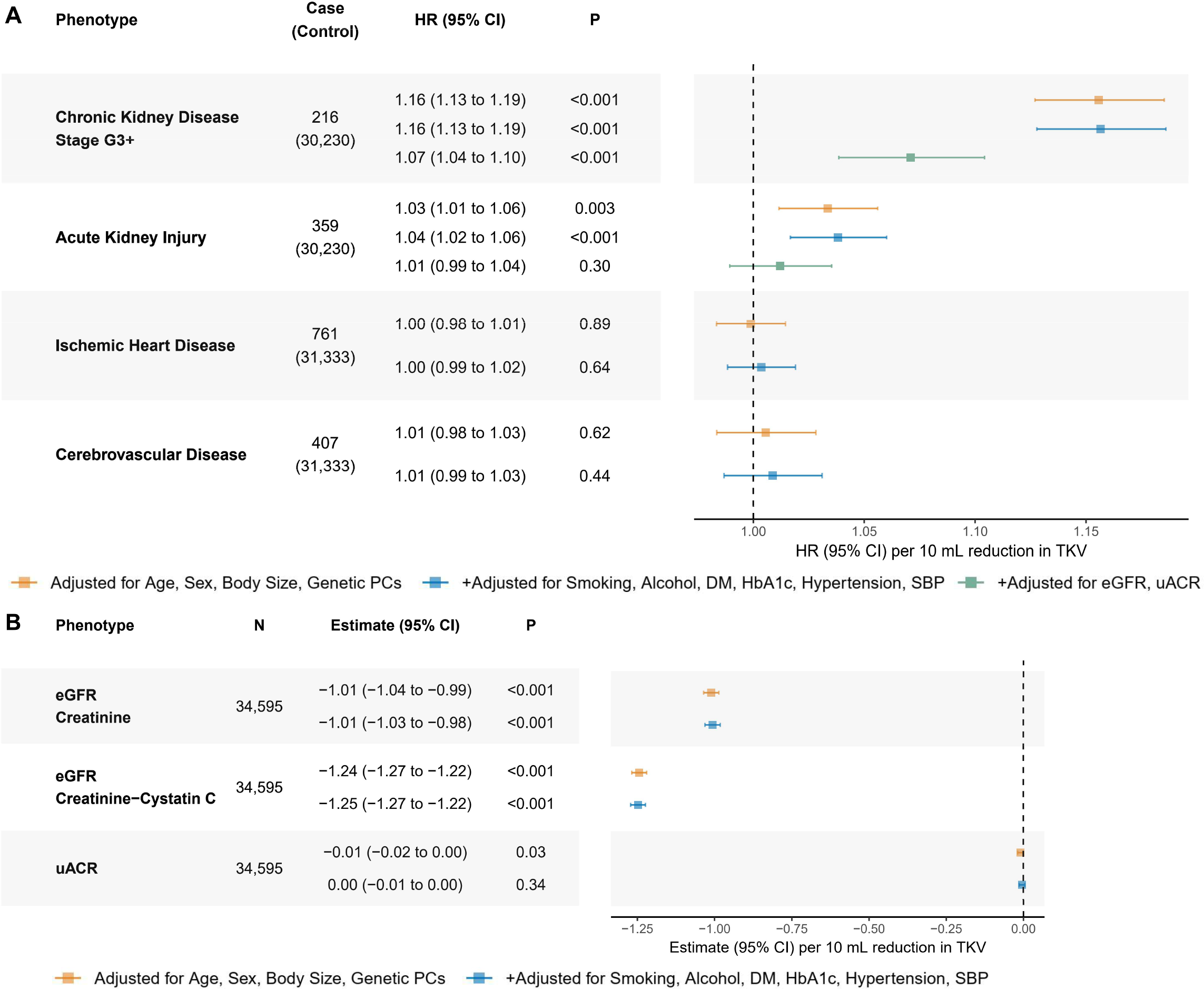
Association of total kidney volume (TKV) with kidney and cardiovascular disease outcomes and measures. Association of TKV with A) incident kidney and cardiovascular outcomes and B) kidney function measures. Models were adjusted for 1) age, sex, body size including body mass index (BMI) and body surface area (BSA), the first 10 genetic principal components of ancestry, image analysis protocol version and assessment center, followed by 2) additionally adjusted for smoking, alcohol, diabetes mellitus (DM), HbA1C, hypertension, and systolic blood pressure (SBP). Association with chronic kidney disease stage G3+ and acute kidney injury were further adjusted for baseline estimated glomerular filtration rate (eGFR) and urine albumin-creatinine ratio (uACR). Significance threshold was corrected for multiple hypotheses testing based on the number of outcomes assessed.

In the second model, further adjusting for risk factors including DM, hypertension, smoking, and alcohol consumption did not alter significance of association with eGFR or incident CKD and AKI events (Figure 1). In the third model, we assessed the association of TKV with incident kidney disease independent of eGFR and uACR. TKV remained significantly associated with incident CKD stage G3+ (HR 1.07, 95% CI 1.04 to 1.10, P < 0.001). However, association with incident AKI was attenuated (HR 1.01, 95% CI 0.99 to 1.04, P = 0.30). Association of TKV with disease diagnoses inclusive of prevalent and incident cases were consistent with that of incident cases only (eFigure 2). We also observed similar results using htTKV and BSA-TKV in lieu of TKV (eFigure 3, 4).

We explored the shape of TKV associations and identified significant nonlinearity with eGFR_CrCys_, CKD stage G3+, and uACR (Figure 2). Smaller TKV was associated with higher risk of CKD stage G3+. However, their association plateaued upon approaching the median. Smaller TKV was also associated with elevated uACR. However, risk of albuminuria increased with larger TKV above the median. We identified nonlinearity with eGFR_Cr_ and incident CKD stage G3+ that were consistent with eGFR_CrCys_ and all cases of CKD stage G3+ (eFigure 5). As with linear analysis, results were similar using htTKV and BSA-TKV in lieu of TKV (eFigure 7, 8). We also investigated nonlinear associations of TKV with ischemic heart and cerebrovascular diseases but did not observe such relationship.

**Figure 2:**
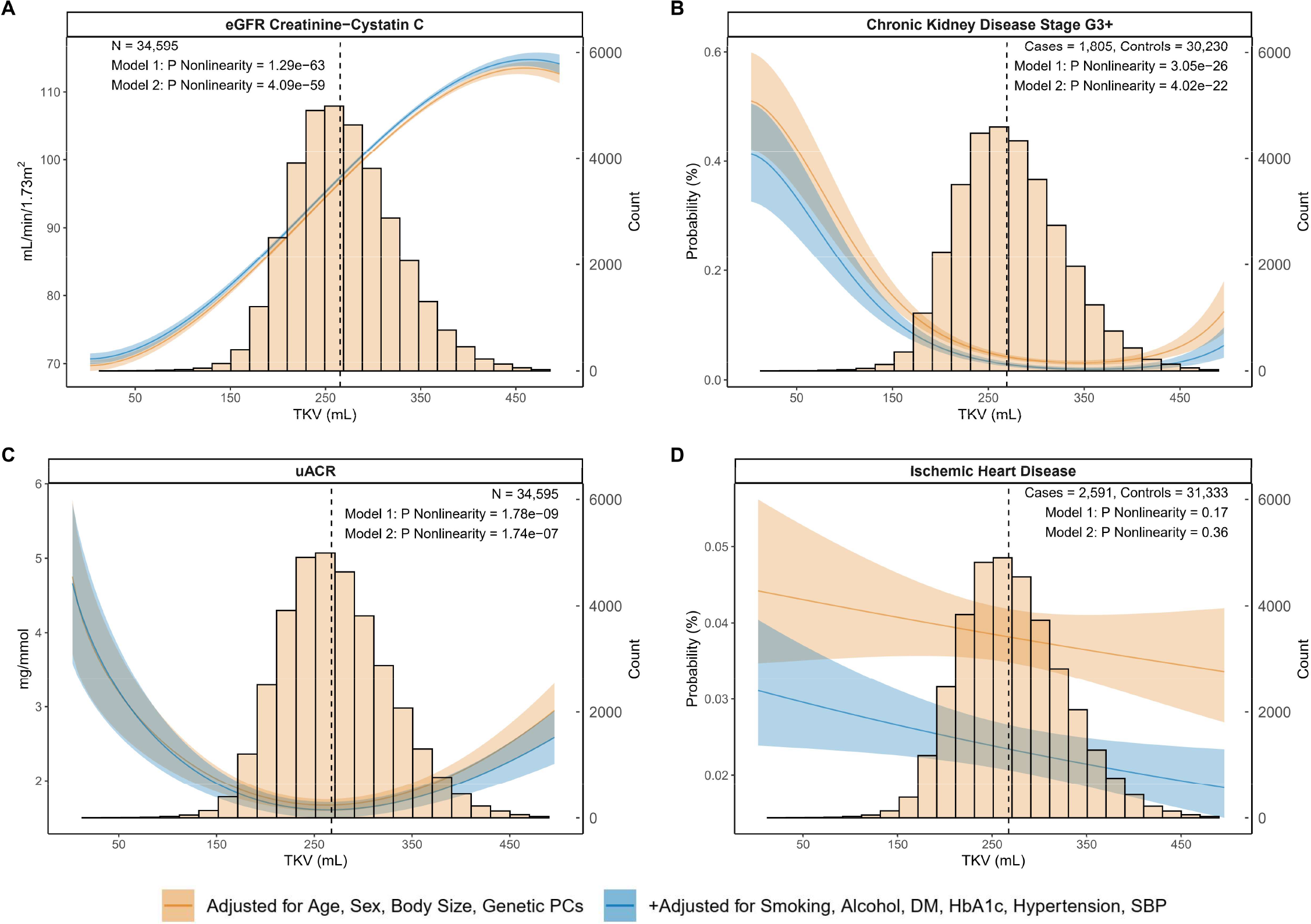
Nonlinear association of total kidney volume (TKV) with kidney and cardiovascular disease outcomes and measures. Nonlinear association of TKV with A) estimated glomerular filtration rate (eGFR), B) chronic kidney disease stage G3+, C) urine albumin-to-creatinine ratio (uACR), and D) ischemic heart disease. Model 1 was adjusted for age, sex, body size including body mass index and body surface area, first 10 principal components of ancestry, image analysis protocol version and assessment center. Model 2 was additionally adjusted for smoking, alcohol, diabetes mellitus (DM), HbA1C, hypertension, and systolic blood pressure (SBP). Dashed line marks the median of each histogram.

Given the nonlinear associations of TKV with CKD, we identified prognostically significant thresholds to inform CKD risk stratification. Using a modified Mazumdar method, thresholds at 185 (4^th^ percentile) and 270 mL (52^nd^ percentile) were identified for TKV, at 113 (4^th^ percentile) and 157 mL/m (49^th^ percentile) for htTKV, and at 119 (11^th^ percentile) and 145 mL/m^2^ (52^nd^ percentile) for BSA-TKV. We assessed improvement in CKD risk prediction when thresholds were added to eGFR and uACR. Of the three TKV measurements, BSA-TKV led to the best prediction performance across likelihood ratio, discrimination, calibration, and reclassification analyses (Table 1, eTable 6). Notably, 228 per every 1,000 individuals would benefit from the inclusion of BSA-TKV for risk assessment based on NARI evaluation. In comparison, inclusion of htTKV and TKV improved reclassification of 104 and 73 per 1,000 individuals, respectively (Table 1). Analysis of differences in C-statistic showed the most significant improvement when BSA-TKV were added to eGFR and uACR (eTable 6). Moreover, predicted risk did not significantly deviate from observed risk based on calibration analysis (eFigure 9). We assessed cumulative incidence and hazard ratios of CKD stage G3+ based on BSA-TKV thresholds (Figure 3). Adjusted for confounding variables and risk factors including eGFR and uACR, individuals below 119 mL/m^2^ were 3.36-fold (95% CI 2.25 to 5.01, P < 0.001), while those between 119 and 145 mL/ m^2^ were 1.96-fold (95% CI 1.30 to 2.96, P = 0.001) more likely to develop CKD stage G3+ relative to those above 145 mL/m^2^. We also assessed differences of hazard ratios between subgroups split by age, sex, or BMI. No significant interactions were found when corrected for multiple comparisons. Individuals <119 mL/m^2^ (11^th^ percentile) and between 119 to 145 mL/m^2^ of BSA-TKV were consistently at higher risk across age, sex, and BMI subgroups (eFigure 10). We performed sensitivity analyses to validate BSA-TKV thresholds. We identified the same thresholds in both univariable and multivariable settings (eFigure 11). 95% confidence intervals derived from bootstrap resampling support the identification of two distinct thresholds as opposed to one. Bias correction of prediction metrics using bootstrap resampling did not substantially alter discrimination, reclassification, and calibration derived from the full cohort (eTable 6, 7). Furthermore, prediction performance using TKV measurements as continuous variables were consistent with results using thresholds (eTable 6, 7).

**Table 1:** Improvement in category free risk reclassification when total kidney volume (TKV), height adjusted TKV (htTKV), or body surface area adjusted TKV (BSA-TKV) thresholds were added to a baseline model of eGFR and uACR, relative to the baseline model without volume thresholds.

| Category Free Net<br>Reclassification Index | %<sup>a</sup> | 95% CI | NARI<sup>b</sup> | P | Bias Corrected % | Bias Corrected NARI |
| --- | --- | --- | --- | --- | --- | --- |
| **TKV Thresholds** |  |  |  |  |  |  |
| **Event (n = 216)** | 0 | -13.34 to 13.34 | 0 | >0.99 | -1.27 | -0.09 |
| **Non-Event (n = 30,230)** | 7.36 | 6.24 to 8.48 | 73.08 | <0.001 | 7.35 | 73.01 |
| **Total (n = 30,446)** | 7.36 | -6.02 to 20.74 | 73.08 | 0.28 | 6.08 | 72.92 |
| **htTKV Thresholds** |  |  |  |  |  |  |
| **Event (n = 216)** | 17.6 | 4.46 to 30.7 | 1.25 | 0.009 | 16.56 | 1.17 |
| **Non-Event (n = 30,230)** | 10.3 | 9.17 to 11.4 | 102.18 | <0.001 | 10.3 | 102.32 |
| **Total (n = 30,446)** | 27.9 | 14.71 to 41.1 | 103.43 | <0.001 | 26.87 | 103.49 |
| **BSA-TKV Thresholds** |  |  |  |  |  |  |
| **Event (n = 216)** | 3.7 | -9.62 to 17.00 | 0.26 | 0.59 | 3.19 | 0.23 |
| **Non-Event (n = 30,230)** | 22.85 | 21.76 to 24.00 | 226.93 | <0.001 | 22.9 | 227.37 |
| **Total (n = 30,446)** | 26.56 | 13.19 to 39.90 | 227.19 | <0.001 | 26.09 | 227.6 |

a. Percentage calculated as difference between number of individuals with increased risk score (if in the event cohort) and those with decreased risk score divided by number of individuals in event, non-event, or total category.
b. Net absolute reclassification index was calculated as the product of reclassification percentage and event rate, multiplied by 1,000. Interpreted as absolute reclassification per 1,000 individuals.

**Figure 3:**
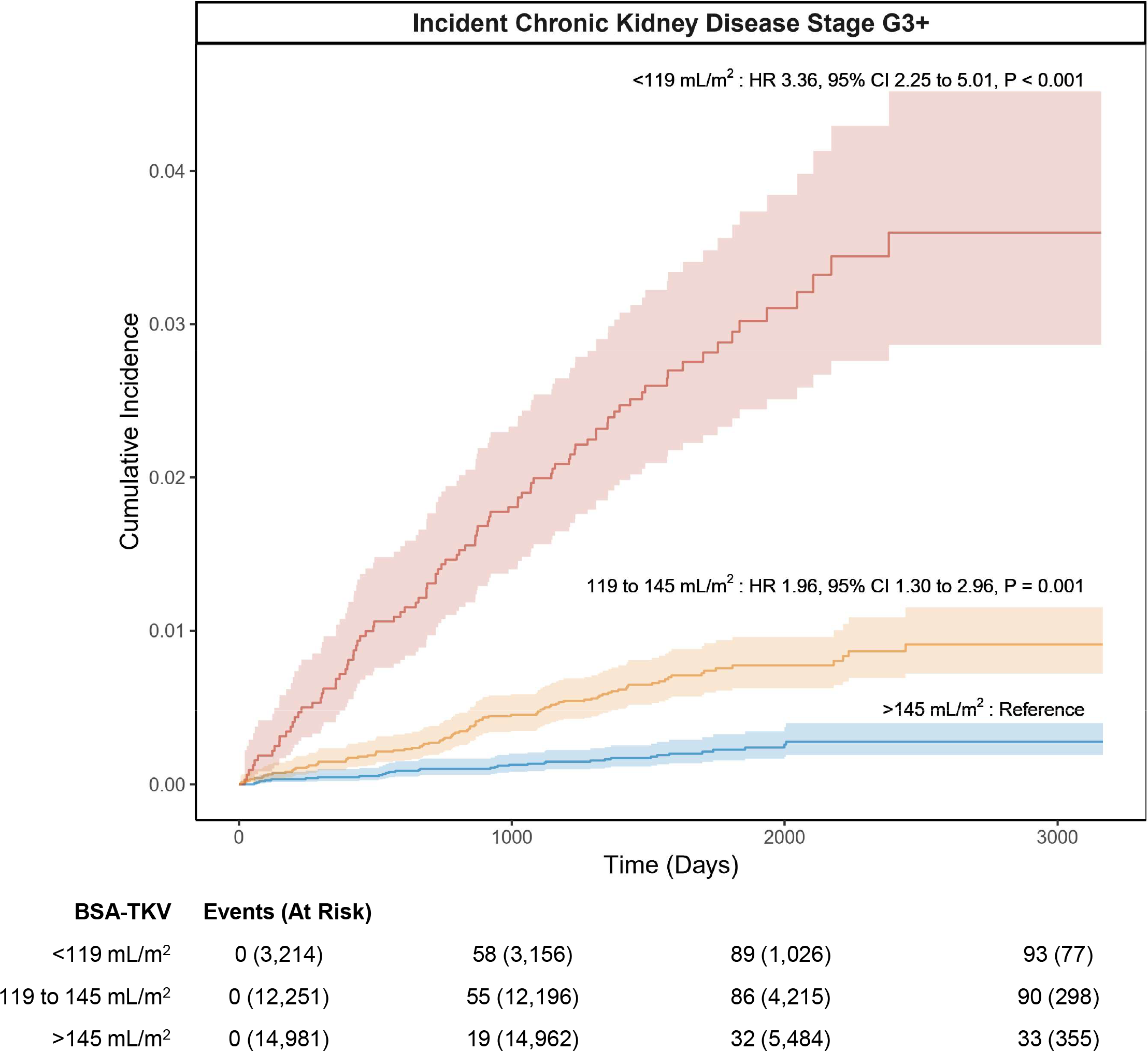
Cumulative incidence and adjusted hazard ratios of incident chronic kidney disease stage G3+ according to body surface area adjusted total kidney volume (BSA-TKV) thresholds. Hazard ratios were adjusted for age, sex, body size including body mass index and body surface area, first 10 genetic principal components of ancestry, smoking, alcohol, diabetes mellitus, HbA1c, hypertension, systolic blood pressure, image analysis protocol version and stratified by assessment center.

We performed two-sample MR using GWAS summary statistics to test the causal association of TKV on kidney and cardiometabolic phenotypes. In the European subset of the CKD Genetics Consortium^22^ (CKDGen) GWAS meta-analysis of eGFR (n = 567,460) and CKD (41,395 cases, 439,303 control participants) independent of UKB, a 10 mL reduction in genetically predicted TKV was associated with 0.28% reduction in eGFR (95% CI -0.46 % to -0.098%, P = 0.002; Figure 4A) and 10% increase in CKD risk (95% CI 4.93% to 15.3%, P < 0.001; Figure 4B). Consistent inverse associations with eGFR were observed using the largest meta-analysis to date by Liu et al.^23^ as well as using the meta-analysis of CKDGen and UKB data sub-grouped by diabetes status (eTable 8).^24^ In contrast, we did not observe significant associations between genetically predicted TKV and the rate of eGFR decline, uACR, coronary artery disease, or stroke (eTable 8). In secondary analyses, we accounted for variant heterogeneity and pleiotropy by excluding outlier variants identified by MR-PRESSO and BMI or BSA associated variants.

**Figure 4:**
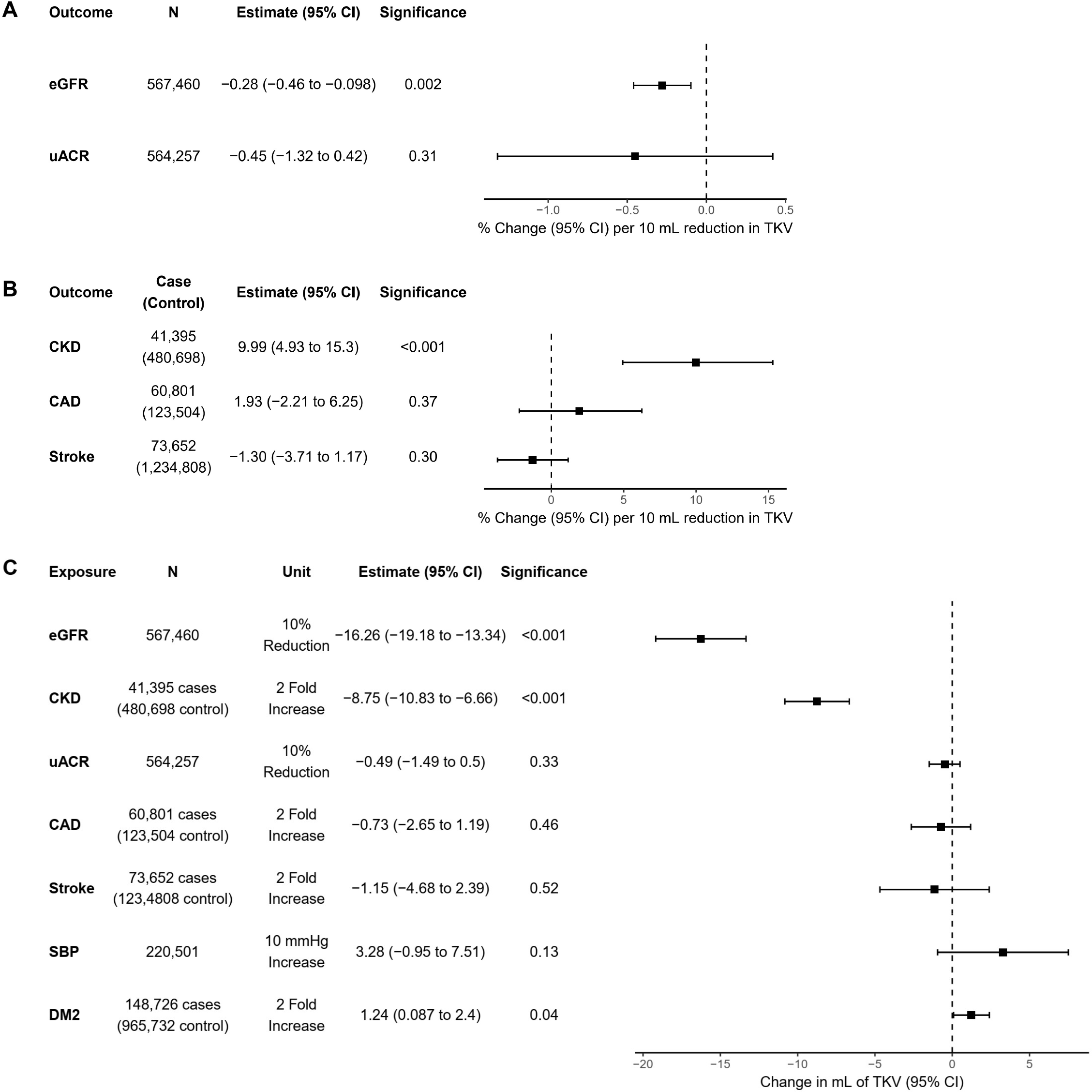
Forward and reverse Mendelian randomization association of genetically predicted total kidney volume (TKV) with kidney and cardiometabolic disease outcomes and measures. Mendelian randomization analysis of genetically predicted TKV as exposure with A) estimated glomerular filtration rate (eGFR), urine albumin-to-creatinine ratio (uACR), B) chronic kidney disease (CKD, defined by eGFR <60 ml/min/1.73m^2^), coronary artery disease (CAD), and stroke as outcomes. C) Reverse Mendelian randomization analysis of genetically predicted phenotypes as exposures and TKV as outcome. In forward MR, estimates are expressed as % change in eGFR, uACR and odds ratio of binary outcomes. Significance threshold was adjusted for multiple hypotheses testing using Bonferroni correction by dividing the number of outcomes assessed in forward or reverse MR analyses respectively.

Results from these analyses were consistent with primary analysis using all SNPs (eTable 9, 10). We also attempted nonlinear MR to discern any significant but nonlinear associations of genetically predicted TKV. However, we were not able to detect significant evidence of nonlinearity potentially due to limitations in event rate per segmented regression blocks (eTable 11).

Next, we tested the reverse effect of genetically predicted eGFR, CKD, and uACR on TKV. A 10% decrease in genetically predicted eGFR based on the European subset of CKDGen GWAS meta-analysis was associated with 16.26 mL reduction in TKV (95% CI -19.18 mL to -13.34 mL, P < 0.001; Figure 4C). Concordantly, a 2-fold increase in risk of genetically predicted CKD was associated with 8.75 mL reduction in TKV (95% CI -10.83 mL to -6.66 mL, P < 0.001). In contrast, uACR was not significantly associated with TKV. We also tested the effect of type 2 diabetes (DM2), SBP, and cardiovascular outcomes on TKV (eTable 12). Among these, a nominally significant association of DM2 was observed (β = 1.24 mL, 95% CI 0.087 to 2.40, P = 0.04) (Figure 4C). Repeat analyses using MR-PRESSO to account for variant heterogeneity and pleiotropy were consistent with primary analysis using all SNPs (eTable 13, 14).

## Discussion

In this study, we demonstrate nonlinear association of kidney volume with eGFR and incident CKD stage G3+. Irrespective of eGFR, uACR and other confounders, a BSA-TKV below the 52^nd^ percentile was associated with a doubling of relative risk for incident CKD stage 3+ and a BSA-TKV below the 11^th^ percentile was associated with a tripling of relative risk for incident CKD stage 3+. These thresholds further translated to improved incident CKD risk prediction beyond eGFR and uACR. Mendelian randomization analysis supports a bidirectional relationship wherein smaller genetically predicted TKV predisposed individuals to higher risk of CKD while higher risk of genetically predicted CKD predisposed individuals to smaller TKV.

Our study utilized the largest available cohort of kidney volume data and affirms TKV as a significant predictor of CKD. We assessed incident kidney and cardiovascular disease development in contrast to previous studies which were limited to a cross-sectional design.^4–6^ Moreover, we observed a nonlinear kidney volume-CKD relationship with exaggerated risk among individuals with kidney volumes below the median. Nonlinear evaluation of kidney volume and function also led to new insights for albuminuria. We found that both larger and smaller kidney volumes were associated with elevated albuminuria, in contrast to previous studies reporting linear or non-significant relationship.^4,5^ Interestingly, we did not find significant association of kidney volume with cardiovascular outcomes. This contrasts the significant association of eGFR and cardiovascular outcomes reported in recent studies.^25,26^

Classification of risk for progression to kidney failure in ADPKD has been revolutionized by the use of age-adjusted htTKV, but empirical assessment comparing TKV, htTKV, and BSA-TKV is lacking.^27,28^ Moreover, past studies of kidney volume did not adjust for eGFR and uACR.^4–6,27,28^ We performed quantitative assessments of TKV, htTKV, and BSA-TKV for CKD risk prediction while adjusting for these traditional biomarkers. These evaluations revealed the superior performance of BSA-TKV when used in conjunction with eGFR and uACR. However, in situations where weight is unavailable, htTKV can also improve CKD risk prediction. Given the improvement upon eGFR and uACR, we identified optimal volume thresholds through a systematic search using a modified Mazumdar method. These thresholds were validated in both univariable and multivariable models to maximize usage potential in settings where covariate data are limited. Our agnostic data-driven method for selecting prognostically significant thresholds is a strength of our study and demonstrates the potential clinical utility of a simple and readily available form of kidney volume for CKD risk stratification.

Radiological and histological assessments have identified atrophy, kidney shrinkage, and glomerulosclerosis as features of CKD.^9,29,30^ However, it was unclear whether a smaller kidney volume caused an increased risk of CKD due to lower nephron endowment or whether kidney volume loss due to atrophy was the result of CKD progression. Mendelian randomization leverages the randomized allocation of alleles at conception to approximate randomized controlled trials and explore the potential causality between exposures and outcomes. As expected, using MR, we demonstrate the relationship between TKV and CKD is the result of both directions of causality, as genetically predicted kidney volume impacts eGFR and CKD risk, and reciprocally, genetically predicted changes in eGFR and CKD impacts TKV.

Our study has important limitations. Observational and MR analyses were limited to individuals of European ancestries. TKV was sourced from a machine learning based method which may differ from traditional methods such as the ellipsoid formula and may not be available in routine practice. Event rate of advanced CKD, particularly end-stage kidney disease, is limited which undermines risk prediction evaluation in vulnerable patient subgroups. We also did not manually evaluate MRI images and as such, cannot rule out the effect of subclinical kidney morphology abnormalities on the associations of TKV. Finally, MR inference of causality is susceptible to confounding by potential sources of pleiotropy.

## Conclusions

We provide observational and genetic evidence in support of TKV as a risk factor for CKD stage G3+. A bidirectional relationship exists whereby smaller kidneys predispose individuals to CKD and CKD leads to smaller kidneys. Risk prediction analyses support the addition of kidney volume to eGFR and uACR for screening advanced CKD.

## Data Availability

All data produced in the present work are available from 1) UK Biobank or 2) GWAS summary statistics from studies listed in supplementary file.

## Author Contributions

Jianhan Wu had full access to all the data in the study and takes responsibility for the integrity of the data and the accuracy of the data analysis.

*Concept and design:* Wu, Lanktree, Paré

*Acquisition, analysis, or interpretation of data:* Wu, Lanktree, Paré

*Drafting of the manuscript:* Wu, Lanktree, Paré

*Critical revision of the manuscript for important intellectual content:* All authors

*Statistical analysis:* Wu

*Obtained funding:* Wu, Lanktree, Paré

*Supervision:* Lanktree, Paré

