## Supplement 1 for "Prognostic Utility of Total Kidney Volume for Chronic Kidney Disease Risk Prediction: An Observational and Mendelian Randomization Study"

**Supplemental Methods**

**Table S1:** Covariate sources and definitions.
**Table S2:** Disease phenotype codes and definitions.
**Table S3:** Genetic variants associated with TKV beyond genome-wide significance.
**Table S4:** Summary of genome-wide association studies used in two-sample Mendelian randomization analyses.
**Table S5:** Baseline demographic characteristics
**Table S6:** Discrimination, calibration, and likelihood ratio test assessing CKD stage G3+ risk prediction performance of eGFR and uACR compared to model incorporating TKV measurements.
**Table S7:** Improvement in category-free risk reclassification when TKV, htTKV, or BSA-TKV were added to a baseline model of eGFR and uACR.
**Table S8:** Inverse variance weighted, weighted median, and MR Egger regression results for each outcome association using all variants.
**Table S9:** Instrument strength, intercept test, and heterogeneity test results for each TKV-outcome association.
**Table S10:** Inverse variance weighted, weighted median, and MR Egger results for each TKV-outcome association, excluding outlier variants identified by MR-PRESSO and BMI or BSA-associated SNPs.
**Table S11:** Nonlinear MR tests of nonlinearity and heterogeneity.
**Table S12:** Inverse variance weighted, weighted median, and MR Egger results for each genetically predicted exposure and TKV association.
**Table S13:** Instrument strength, intercept test, and heterogeneity test results for each exposure-outcome association.
**Table S14:** Inverse variance weighted, weighted median, and MR Egger results for each genetically predicted exposure and TKV association, excluding outlier variants identified by MR-PRESSO.
**Figure S1:** Histogram of TKV, htTKV, or BSA-TKV.
**Figure S2:** Association of TKV with all cases of kidney and cardiovascular disease outcomes.
**Figure S3:** Association of htTKV with kidney and cardiovascular disease outcomes and measures.
**Figure S4:** Association of BSA-TKV with kidney and cardiovascular disease outcomes and measures.
**Figure S5:** Nonlinear association of TKV with $eGFR_{Cr}$ and incident cases of CKD stage G3+.
**Figure S6:** Nonlinear association of htTKV with kidney and cardiovascular disease outcomes and measures
**Figure S7:** Nonlinear association of BSA-TKV with kidney and cardiovascular disease outcomes and measures.
**Figure S8:** Calibration plot comparing observed and predicted risk when BSA-TKV is incorporated in a baseline model with eGFR and uACR.
**Figure S9:** Association of BSA-TKV thresholds with incident CKD stage G3+ split by age, sex, and BMI subgroups.
**Figure S10:** Histogram and summary statistics of BSA-TKV thresholds identified from 1,000 iterations of bootstrap resampling in three different univariable and multivariable settings.

### Supplemental Methods

eGFR and uACR Assessment in UK Biobank

Race-free eGFR (mL/min/m$^2$) was calculated using serum creatinine (mg/dL) and cystatin C (mg/L) according to the 2021 CKD-EPI creatinine ($eGFR_{Cr}$) and creatinine-cystatin C ($eGFR_{CrCys}$) equations. uACR (mg/mmol) was calculated by dividing urine microalbumin (mg/L) and urine creatinine (mmol/L). Biochemistry and urine data were winsorized according to reportability limits. Individuals with undetectable urine albumin were assigned the lower limit of detection (6.7 mg/L).

Covariates

Participants' age, alcohol consumption, and smoking status were collected from self-report questionnaire. Image processing protocol version and assessment centre were included as nuisance factors. Genetic sex and genetic principal components (PC) were obtained from genotype data. Height (m), weight (kg), and BMI (kg/m$^2$) were collected from physical measurements. Missing height and weight were assigned values from overlapping data fields in UK Biobank (Table S1).

Systolic blood pressure (SBP), hypertension, HbA1c and diabetes mellitus (DM) were also included as additional covariates. SBP (mmHg) measurements were amalgamated from both manual and automated readings. HbA1c (mmol/mol) was collected from blood biochemistry data and winsorized according to reportability limits. HbA1c mmol/mol was converted to % using the conversion factor (1/10.929) + 2.15. Hypertension was defined by ICD10 phecode 401 (Table S2). DM was defined by ICD10 phecode 250.

Disease Outcome Assessment

Disease outcomes were assessed according to eTable 2. Incident cases and time-to-event were defined based on time of imaging. Analyses of incident chronic kidney disease (CKD) stage G3+ and acute kidney injury (AKI) excluded individuals with any baseline eGFR < 60 mL/min/m$^2$, uACR > 30 mg/mmol, or records of CKD stage G3+ and AKI prior to time of imaging. Analyses of incident cardiovascular outcomes excluded individuals who meet any of the listed definitions for ischemic heart or cerebrovascular diseases prior to time of imaging. Control groups for CKD stage G3+ and AKI excluded individuals with any codes for CKD (ICD10 N18.0 to N18.5), AKI (ICD10 N17.0 to N17.9), dialysis (ICD10 Y60.2, Y84.1, Z49.0, Z49.1, Z49.2, Z99.2; OPCS4 X40 to X42), algorithmically defined end-stage kidney disease (UKB FID 42027), eGFR <60 mL/min/m$^2$, or uACR >= 3 mg/mmol. Control groups for cardiovascular outcomes excluded individuals who meet any of the listed definitions for ischemic heart and cerebrovascular disease.

Nonlinear Association Analyses

TKV measurements underwent nonlinearity testing and appropriate transformation using the multivariate fractional polynomial method (R mfp package v 1.5.2.2).[1] Continuous covariates also underwent nonlinearity testing and transformation to optimize model adjustment accuracy. This method involves an iterative selection of the best-fitting model across a series of fractional polynomial transformations, comparing to the linear model or the previous model of best fit as the null hypothesis. The optimal transformation was selected based on 1) significance of nonlinearity followed by 2) significance of improvement using 2$^{nd}$-degree polynomial compared to 1$^{st}$-degree polynomial transformations.

Threshold Identification

We utilized a modified Mazumdar[2,3] method by expanding the search for optimal thresholds beyond a single point and by utilizing bootstrap resampling to validate distinct and significant thresholds. Thresholds were identified for each of TKV, htTKV, and BSA-TKV by iteratively evaluating all integers by increment of 1 unit across the 2.5$^{th}$ to 97.5$^{th}$ percentile of each TKV measurement. The first threshold was selected based on the significance of likelihood ratio test compared to a model without any threshold. The second threshold was assessed based on significance of likelihood ratio compared to a model with one threshold. This process was repeated 1,000 times using bootstrap resampling (R boot package v 1.3.28.1) to calculate bias, standard deviation, and frequency. We considered thresholds to be significant and distinct if they fall outside of the 95% confidence intervals of the other threshold (Figure S10).

Two Sample Mendelian Randomization

Independent genetic variants of outcomes and exposures at genome-wide significance ($P < 5 \times 10^{-8}$) were pruned based on linkage disequilibrium at a threshold of $r^2 < 0.001$ using the European subset of the 1000 Genomes phase 3 panel as the reference.[4]

Weighted-median, inverse-variance weighted, MR-Egger, heterogeneity, and intercept tests were performed using the R TwoSampleMR package (v 0.5.6). Directional pleiotropy was ascertained based on the significance ($P < 0.05$) of Egger intercept deviation from 0.[5] MR-PRESSO was performed using the R MRPRESSO package (v 1.0). MR-PRESSO first assesses the presence of overall horizontal pleiotropy within a set of genetic instruments based on the significance of global test p-value ($P < 0.05$) and using 10,000 simulations.[6] Subsequently, outlier and invalid variants with potential pleiotropic effects were identified and removed should the global test p-value reach significance. Instrument strength were estimated based on $R^2$ and F-statistic.

In forward MR, regression coefficients were converted from per 1 standard deviation (SD) increase in TKV to per 10 mL decrease in TKV by first multiplying -1, then dividing by 1 SD of TKV (unit in L) and multiplying by 10. In reverse MR, coefficients for eGFR and uACR were converted from per 1 logarithmic unit increase to per 10% reduction by multiplying -1 and the natural logarithm of 1.1. Coefficients for binary outcomes were converted from per 1 log odd increase to per 2-fold increase by multiplying the natural logarithm of 2.

Nonlinear Mendelian Randomization

Nonlinear Mendelian randomization (MR) analyses were performed using the R nlmr[7] package (v 2.0). Residual TKV was calculated by subtracting genetically predicted TKV from observed TKV. Residual TKV was then stratified into deciles and localized average causal effects (LACE) were estimated while adjusting for age, sex, assessment centre, image processing protocol version, and the first 10 genetic PCs. Nonlinearity across LACE estimates were assessed using the fractional polynomial (FP), quadratic, and Cochran Q test.[7] Validity of nonlinear MR result relies on the assumption of a constant instrumental variable and exposure relationship. Heterogeneity, or non-constant association between instrumental variable and exposure, were assessed using the Q test and quadratic trend test of heterogeneity (Table S11).

**Figure S1:** Histogram of TKV, htTKV, or BSA-TKV.

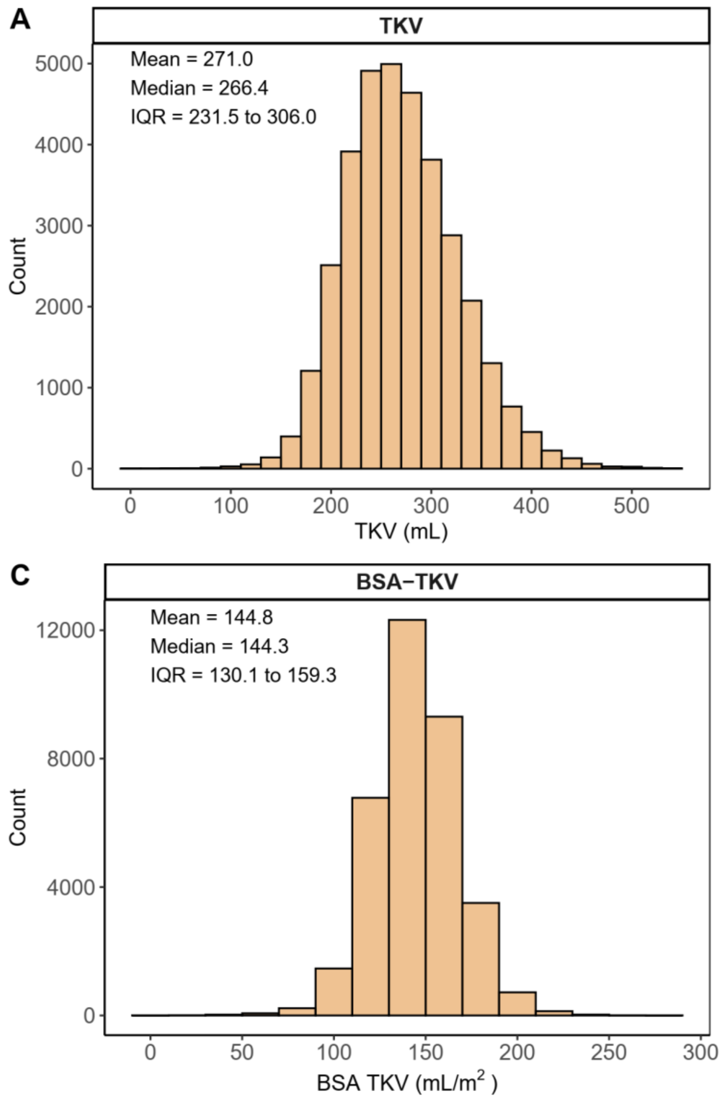

**Figure S2:** Association of TKV with all cases of kidney and cardiovascular disease outcomes.

| Phenotype | Case (Control) | OR (95% CI) | P |
|---|---|---|---|
| **Chronic Kidney Disease Stage G3+** | 1,805<br>(30,230) | 1.07 (1.06 to 1.08)<br>1.08 (1.06 to 1.09)<br>1.03 (1.01 to 1.05) | <0.001<br><0.001<br>0.006 |
| **Acute Kidney Injury** | 499<br>(30,230) | 1.06 (1.04 to 1.08)<br>1.06 (1.04 to 1.08)<br>1.03 (1.01 to 1.05) | <0.001<br><0.001<br>0.007 |
| **Albuminuria** | 2,859<br>(29,572) | 0.98 (0.97 to 0.99)<br>0.98 (0.97 to 0.99) | <0.001<br>0.002 |
| **Ischemic Heart Disease** | 2,591<br>(31,333) | 1.01 (1.00 to 1.01)<br>1.01 (1.00 to 1.02) | 0.21<br>0.02 |
| **Cerebrovascular Disease** | 842<br>(31,333) | 1.01 (1.00 to 1.03)<br>1.01 (1.00 to 1.03) | 0.16<br>0.09 |

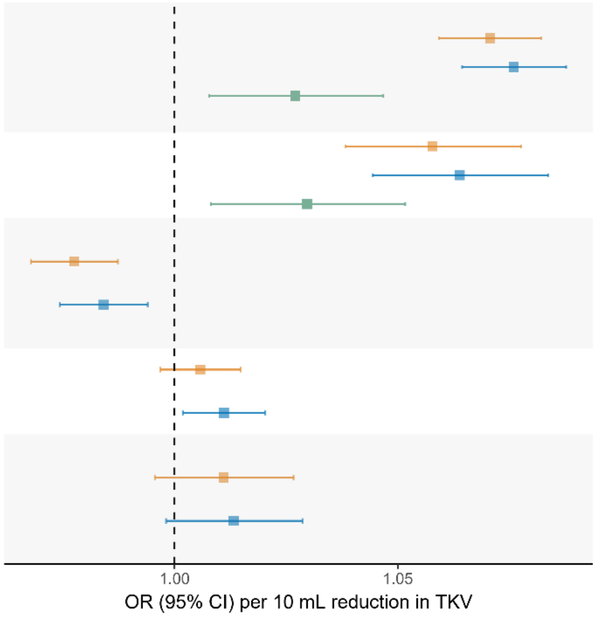


Adjusted for Age, Sex, Body Size, Genetic PCs — +Adjusted for Smoking, Alcohol, DM, HbA1c, Hypertension, SBP — +Adjusted for eGFR, uACR

Association of TKV with all cases of disease outcomes were adjusted for 1) age, sex, body size including BMI and BSA, the first 10 genetic PCs of ancestry, image analysis protocol version and assessment centre, followed by 2) additionally adjusted for smoking, alcohol, DM, HbA1C, hypertension, and SBP. Association with CKD stage G3+ and AKI were further adjusted for eGFR and uACR. Significance threshold was corrected for multiple hypotheses testing based on the number of outcomes assessed.

**Figure S3:** Association of htTKV with kidney and cardiovascular disease outcomes and measures.

**A**

| Phenotype | Case (Control) | HR (95% CI) | P |
|---|---|---|---|
| **Chronic Kidney Disease Stage G3+** | 216 (30,230) | 1.65 (1.51 to 1.80) | <0.001 |
| | | 1.65 (1.52 to 1.80) | <0.001 |
| | | 1.26 (1.14 to 1.40) | <0.001 |
| **Acute Kidney Injury** | 359 (30,230) | 1.12 (1.04 to 1.21) | 0.002 |
| | | 1.14 (1.06 to 1.22) | <0.001 |
| | | 1.04 (0.96 to 1.13) | 0.30 |
| **Ischemic Heart Disease** | 761 (31,333) | 1.00 (0.94 to 1.05) | 0.88 |
| | | 1.01 (0.96 to 1.07) | 0.64 |
| **Cerebrovascular Disease** | 407 (31,333) | 1.02 (0.94 to 1.10) | 0.68 |
| | | 1.03 (0.95 to 1.11) | 0.48 |

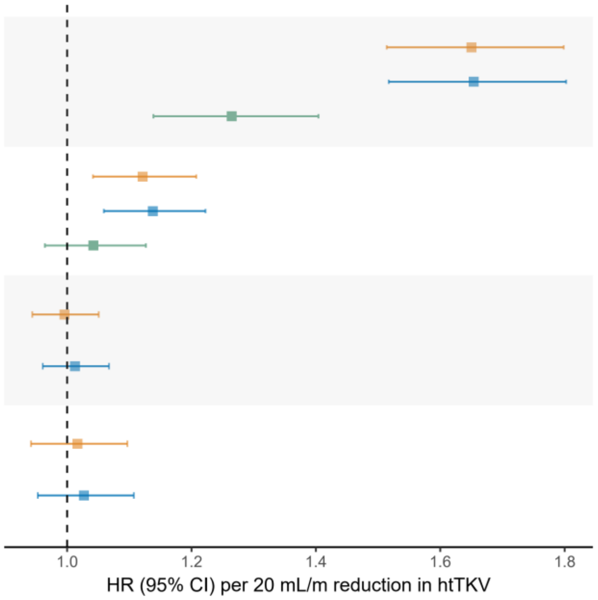


Adjusted for Age, Sex, Body Size, Genetic PCs; +Adjusted for Smoking, Alcohol, DM, HbA1c, Hypertension, SBP; +Adjusted for eGFR, uACR

**B**

| Phenotype | N | Estimate (95% CI) | P |
|---|---|---|---|
| **eGFR Creatinine** | 34,595 | −3.47 (−3.55 to −3.38) | <0.001 |
| | | −3.45 (−3.54 to −3.37) | <0.001 |
| **eGFR Creatinine−Cystatin C** | 34,595 | −4.27 (−4.35 to −4.19) | <0.001 |
| | | −4.28 (−4.37 to −4.20) | <0.001 |
| **uACR** | 34,595 | −0.03 (−0.06 to 0.00) | 0.04 |
| | | −0.02 (−0.05 to 0.01) | 0.25 |

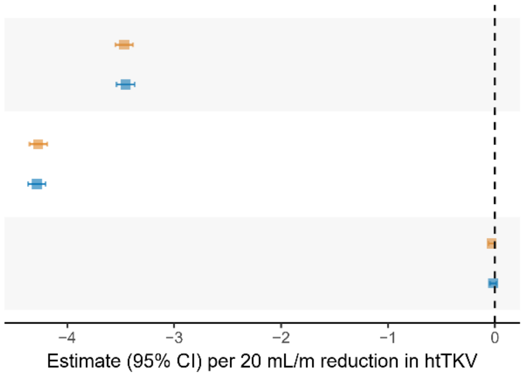


Adjusted for Age, Sex, Body Size, Genetic PCs; +Adjusted for Smoking, Alcohol, DM, HbA1c, Hypertension, SBP

Association of htTKV with A) incident outcomes and B) kidney function biomarkers. Models were adjusted for 1) age, sex, body size including BMI and BSA, the first 10 genetic PCs, and nuisance factors including image analysis protocol version and assessment centre, followed by 2) additionally adjusted for smoking, alcohol, DM, HbA1C, hypertension, and SBP. Association with CKD stage G3+ and AKI were further adjusted for eGFR and uACR. Significance threshold was corrected for multiple hypotheses testing based on the number of outcomes assessed.

**Figure S4:** Association of BSA-TKV with kidney and cardiovascular disease outcomes and measures

**A**

| Phenotype | Case (Control) | HR (95% CI) | P |
|---|---|---|---|
| Chronic Kidney Disease Stage G3+ | 216 (30,230) | 1.82 (1.65 to 2.00) | <0.001 |
| | | 1.82 (1.65 to 2.00) | <0.001 |
| | | 1.34 (1.19 to 1.52) | <0.001 |
| Acute Kidney Injury | 359 (30,230) | 1.15 (1.05 to 1.25) | 0.002 |
| | | 1.17 (1.07 to 1.27) | <0.001 |
| | | 1.05 (0.96 to 1.16) | 0.26 |
| Ischemic Heart Disease | 761 (31,333) | 1.00 (0.94 to 1.06) | 0.95 |
| | | 1.02 (0.96 to 1.08) | 0.59 |
| Cerebrovascular Disease | 407 (31,333) | 1.02 (0.93 to 1.11) | 0.72 |
| | | 1.03 (0.94 to 1.12) | 0.53 |

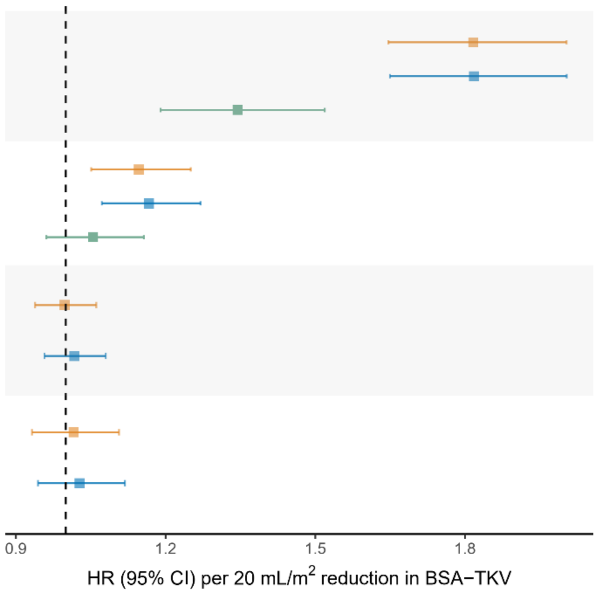


Adjusted for Age, Sex, Body Size, Genetic PCs — +Adjusted for Smoking, Alcohol, DM, HbA1c, Hypertension, SBP — +Adjusted for eGFR, uACR

**B**

| Phenotype | N | Estimate (95% CI) | P |
|---|---|---|---|
| eGFR Creatinine | 34,595 | −3.90 (−4.00 to −3.81) | <0.001 |
| | | −3.89 (−3.98 to −3.79) | <0.001 |
| eGFR Creatinine−Cystatin C | 34,595 | −4.81 (−4.90 to −4.72) | <0.001 |
| | | −4.83 (−4.92 to −4.74) | <0.001 |
| uACR | 34,595 | −0.04 (−0.07 to 0.00) | 0.04 |
| | | −0.02 (−0.05 to 0.01) | 0.26 |

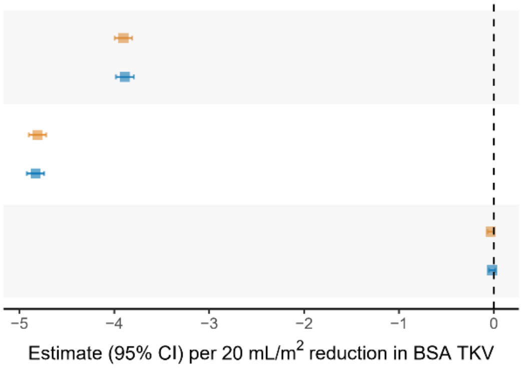


Adjusted for Age, Sex, Body Size, Genetic PCs — +Adjusted for Smoking, Alcohol, DM, HbA1c, Hypertension, SBP

Association of BSA-TKV with A) incident outcomes and B) kidney function biomarkers. Models were adjusted for 1) age, sex, body size including BMI and BSA, the first 10 genetic PCs, and nuisance factors including image analysis protocol version and assessment centre, followed by 2) additionally adjusted for smoking, alcohol, DM, HbA1C, hypertension, and SBP. Association with CKD stage G3+ and AKI were further adjusted for eGFR and uACR. Significance threshold was corrected for multiple hypotheses testing based on the number of outcomes assessed.

**Figure S5:** Nonlinear association of TKV with $eGFR_{Cr}$ and incident cases of CKD stage G3+.

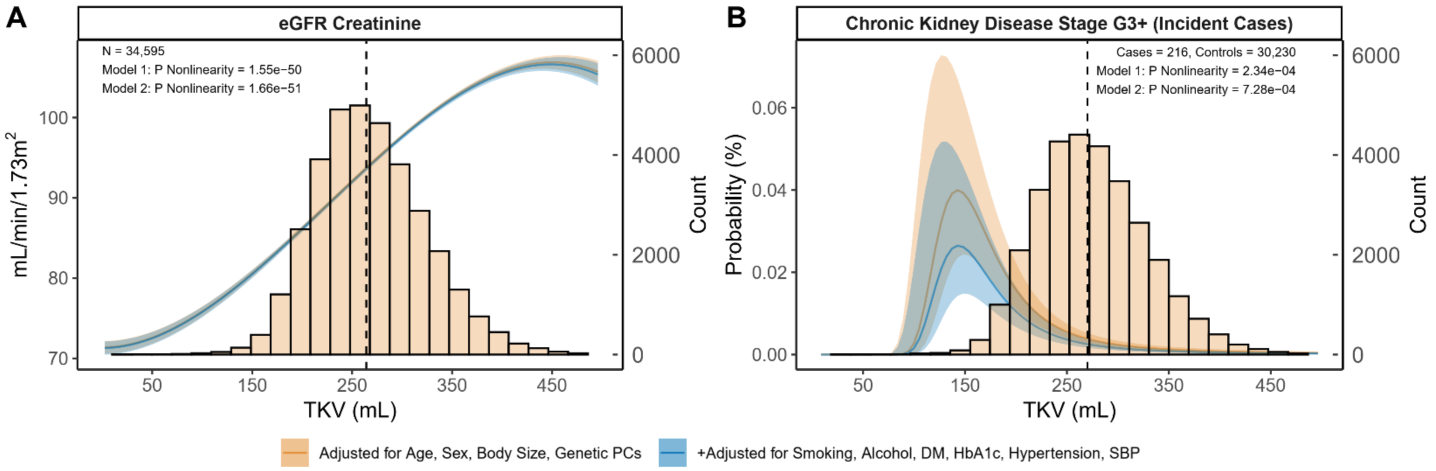


Model 1 was adjusted for age, sex, body size including BMI and BSA, and first 10 PCs. Model 2 was additionally adjusted for smoking, alcohol, DM, HbA1C, hypertension, and SBP. Dashed line marks the median of each histogram.

**Figure S6:** Nonlinear association of htTKV with kidney and cardiovascular disease outcomes and measures.

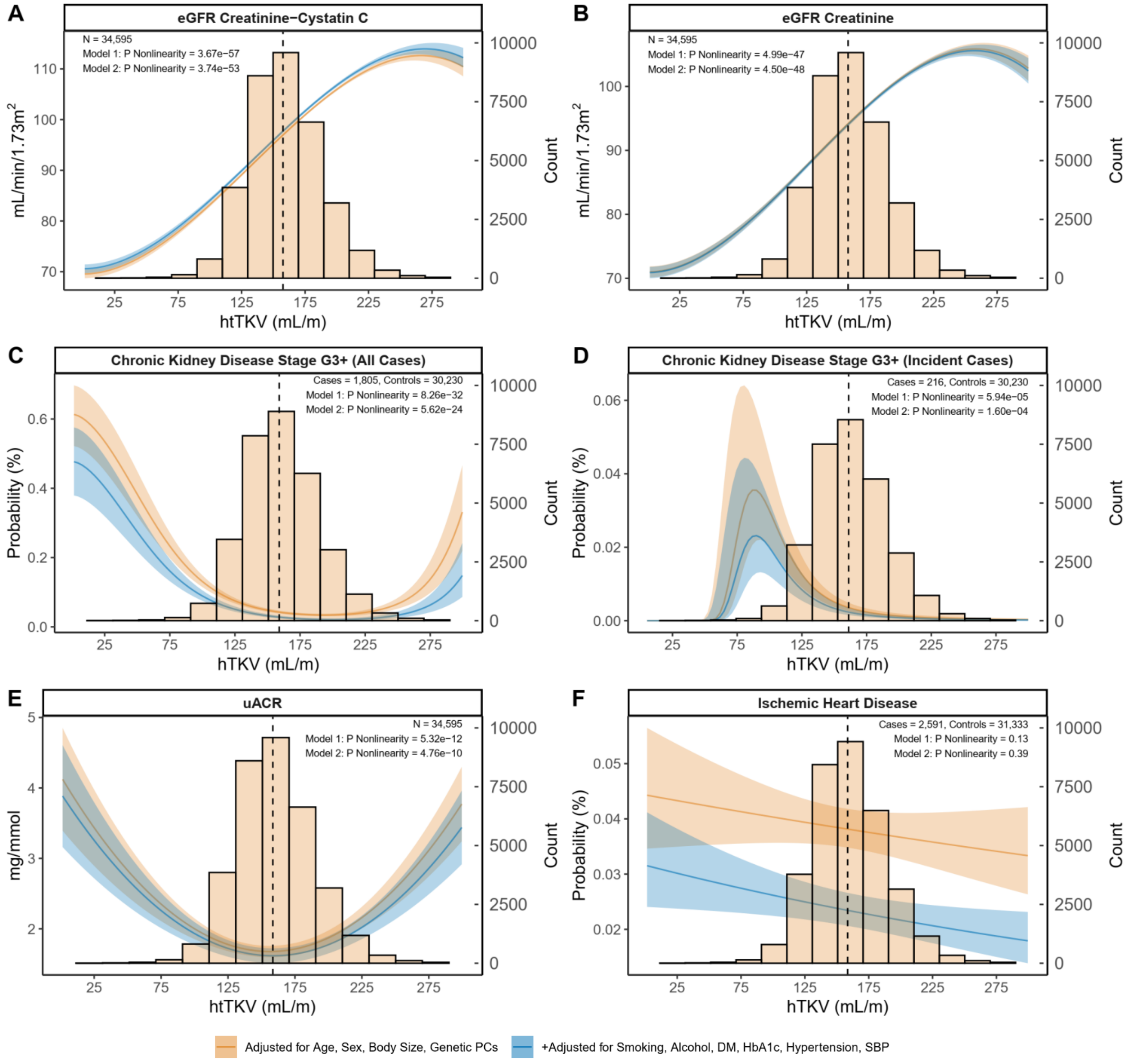


Model 1 was adjusted for age, sex, body size including BMI and BSA, and first 10 PCs. Model 2 was additionally adjusted for smoking, alcohol, DM, HbA1C, hypertension, and SBP. Dashed line marks the median of each histogram.

**Figure S7:** Nonlinear association of BSA-TKV with kidney and cardiovascular disease outcomes and measures.

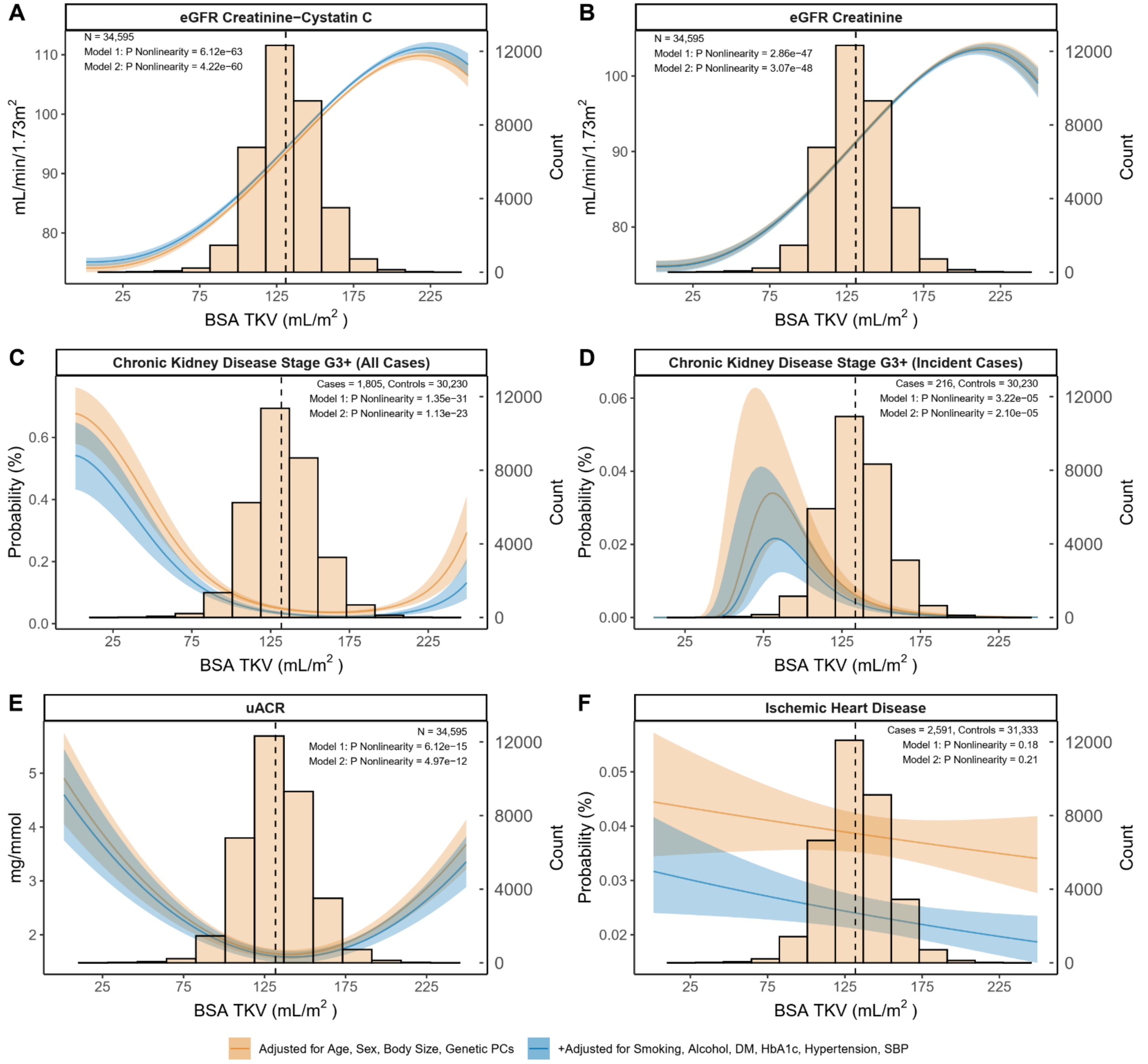


Model 1 was adjusted for age, sex, body size including BMI and BSA, and first 10 PCs. Model 2 was additionally adjusted for smoking, alcohol, DM, HbA1C, hypertension, and SBP. Dashed line marks the median of each histogram.

**Figure S8:** Calibration plot comparing observed and predicted risk when BSA-TKV is incorporated in a baseline model with eGFR and uACR.

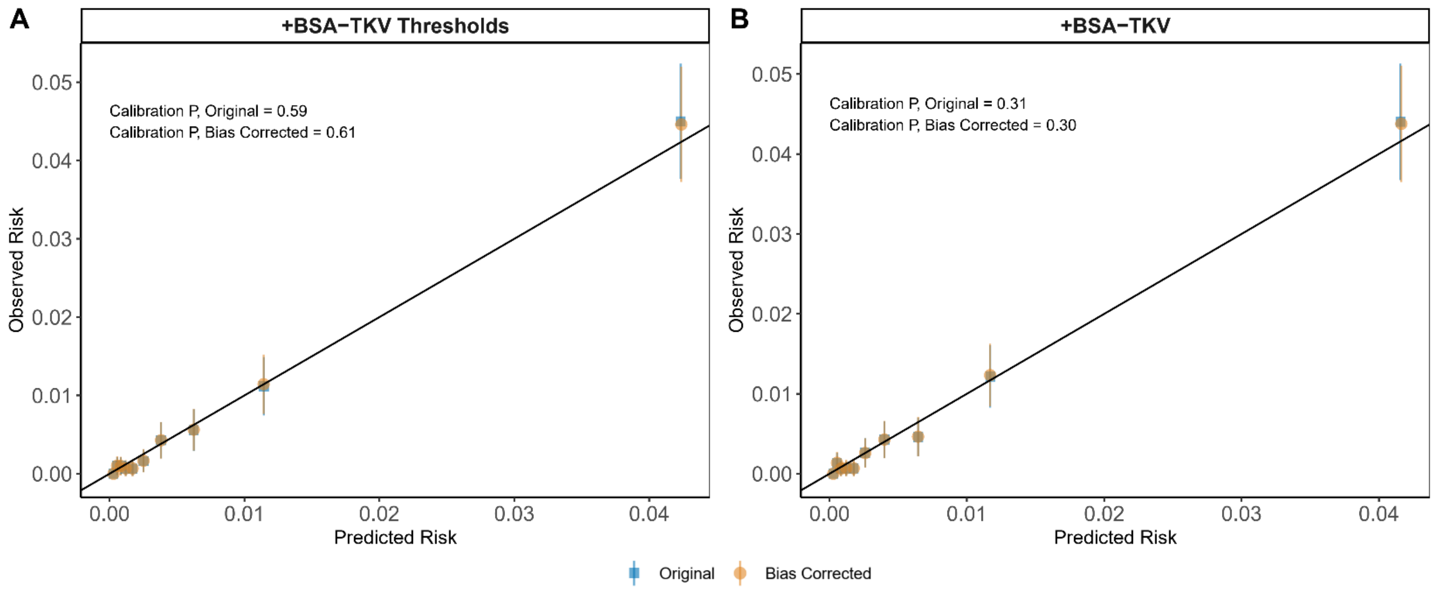


Significance of differences between predicted and observed risk was assessed using the Hosmer-Lemeshow test. Bias was corrected using Harrell's bias correction method.

**Figure S9:** Association of BSA-TKV thresholds with incident CKD stage G3+ split by age, sex, and BMI subgroups.

| Category | Case (Control) | HR (95% CI) | P | Threshold | Subgroup | P Interaction |
|---|---|---|---|---|---|---|
| Sex | 54 (2,036) | 4.48 (2.11 to 9.52) | <0.001 | <119 mL/m$^2$ | Female | 0.13 |
| | 39 (1,085) | 4 (2.25 to 7.12) | <0.001 | | Male | |
| | 44 (6,924) | 2.47 (1.18 to 5.14) | 0.02 | 119 to 145 mL/m$^2$ | Female | 0.73 |
| | 46 (5,237) | 1.78 (1.06 to 2.99) | 0.03 | | Male | |
| BMI | 70 (2,414) | 4.39 (2.59 to 7.44) | <0.001 | <119 mL/m$^2$ | BMI < 30 | 0.21 |
| | 23 (707) | 3.2 (1.37 to 7.49) | 0.007 | | BMI >= 30 | |
| | 71 (10,107) | 2.11 (1.3 to 3.44) | 0.003 | 119 to 145 mL/m$^2$ | BMI < 30 | 0.59 |
| | 19 (2,054) | 1.62 (0.73 to 3.61) | 0.2 | | BMI >= 30 | |
| Age | 13 (1,250) | 5.54 (1.77 to 17.28) | 0.003 | <119 mL/m$^2$ | Age < 65 | 0.10 |
| | 80 (1,871) | 3.49 (2.15 to 5.64) | <0.001 | | Age >= 65 | |
| | 23 (5,967) | 4.21 (1.53 to 11.56) | 0.005 | 119 to 145 mL/m$^2$ | Age < 65 | 0.02 |
| | 67 (6,194) | 1.62 (1.03 to 2.57) | 0.04 | | Age >= 65 | |

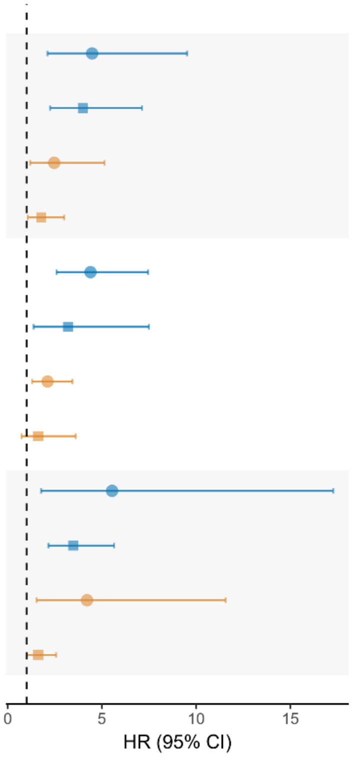


Analysis was adjusted for age, sex, body size including BMI and BSA, first 10 PCs, smoking, alcohol, DM, HbA1C, hypertension, SBP, eGFR and uACR. Significance threshold was adjusted for multiple hypotheses testing using Bonferroni correction based on the number of subgroups assessed.

**Figure S10:** Histogram and summary statistics of BSA-TKV thresholds identified from 1,000 iterations of bootstrap resampling.

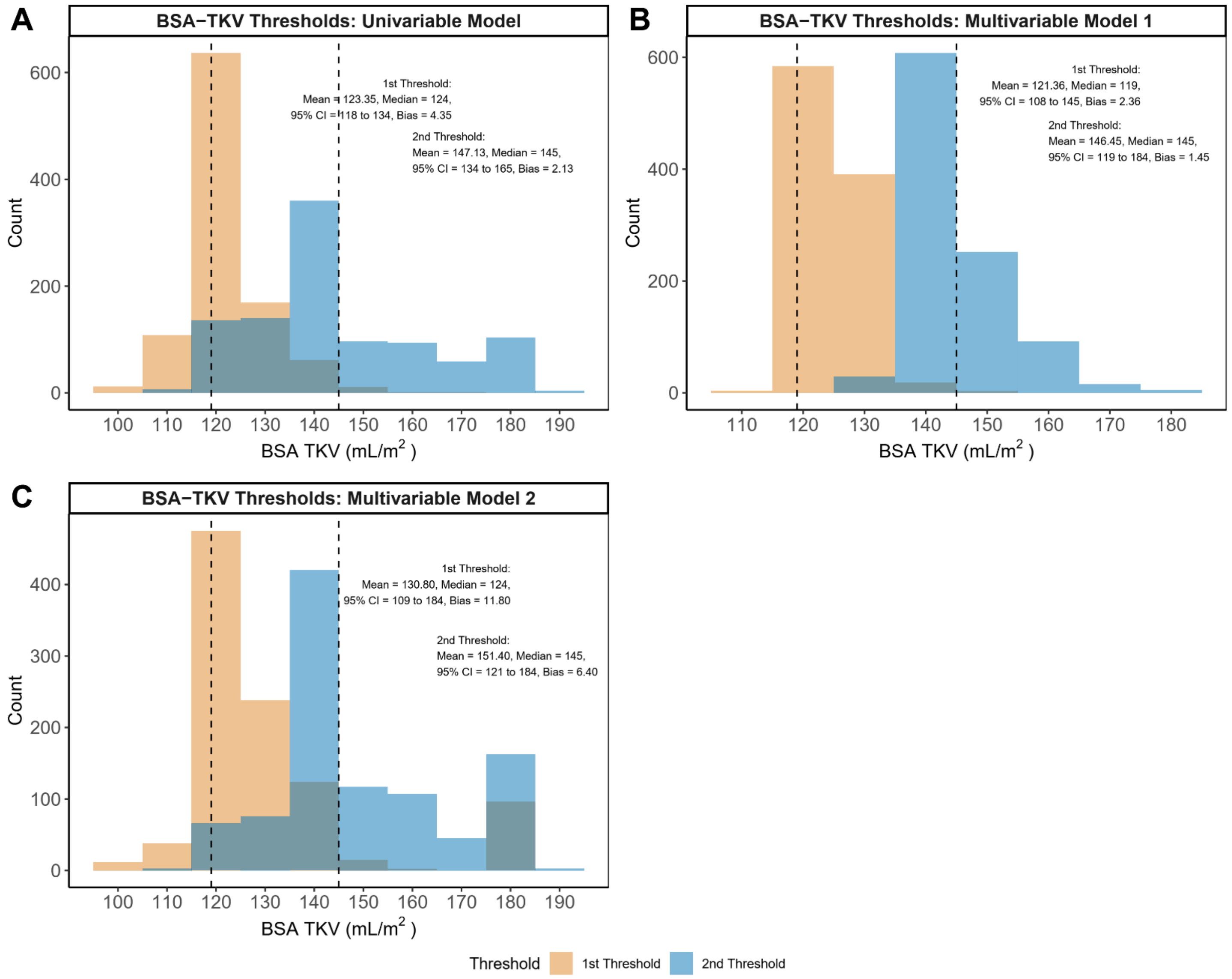


Thresholds were identified in A) a univariable model consisting of only BSA-TKV thresholds, B) a multivariate model consisting of BSA-TKV thresholds in addition to eGFR and uACR, C) a multivariate model consisting of BSA-TKV thresholds, eGFR, uACR, age, sex, body size, smoking, alcohol, diabetes, HBA1c, hypertension, SBP, genetic PCs, analysis protocol, and stratified by assessment center. Dashed lines represent thresholds derived from the full cohort.

**Table S1:** Covariate sources and definitions.

| Phenotype | FID | Category | Definition |
|---|---|---|---|
| Left Kidney Volume | 21081 | 158 | Left kidney volume |
| Right Kidney Volume | 21082 | 158 | Right kidney volume |
| Total Kidney Volume | NA | NA | Sum of left and right kidney volumes |
| Height Standardized Total Kidney Volume | NA | NA | Total kidney volume divided by height |
| Body Surface Area Standardized Total Kidney Volume | NA | NA | Total kidney volume divided by body surface area |
| IDP Pipeline Version | 21094 | 158 | Analysis protocol versions used to derive kidney volume |
| Assessment Centre | 54 | 100113 | Participant assessment center for UK Biobank recruitment |
| Age | 21003 | 1001 | Age of participants at time of assessment |
| Genetic Sex | 22001 | 100313 | Genetic sex determined by genotyping analysis |
| Genetic Principal Components | 22009 | 716 | Principal component scores derived from genotyping analysis |
| Smoking Status | 20116 | 704 | Self-reported current or past smoking status at time of assessment |
| Alcohol Intake Frequency | 1558 | 704 | Self-reported alcohol intake frequency at time of assessment |
| Systolic Blood Pressure | 4080, 93 | 706 | Two measures of blood pressure taken a few moments apart using either an automated machine or manual sphygometer |
| Height | 50, 12144 | 706 | Standing height at time of assessment |
| Weight | 3160, 21002, 23098 | 706 | Body weight at time of assessment |
| BMI | 21001 | 706 | BMI at time of assessment |
| BSA | NA | NA | Body surface area based on the DuBois equation at time of assessment |
| HBA1c | 30750 | 717 | Hemoglobin A1c at time of initial, or if unavailable, follow-up assessment |
| Creatinine | 30700 | 717 | Creatinine at time of initial, or if unavailable, follow-up assessment |
| Cystatin C | 30720 | 717 | Cystatin C at time of initial, or if unavailable, follow-up assessment |
| eGFR Creatinine | NA | NA | eGFR creatinine calculated using CKD-EPI Creatinine Equation 2021 |
| eGFR Creatinine and Cystatin C | NA | NA | eGFR creatinine calculated using CKD-EPI Creatinine-Cystatin Equation 2021 |
| Urine Creatinine | 30510 | 100083 | Enzymatic creatinine in urine at time of initial, or if unavailable, follow-up assessment |
| Urine Microalbumin | 30500 | 100083 | Microalbumin in urine at time of initial, or if unavailable, follow-up assessment |
| uACR | NA | NA | Urine albumin-creatinine ratio calculated as ratio of urine microalbumin and urine creatinine |

**Table S2:** Disease phenotype codes and definitions.

| Disease Phenotype | Code (ICD10, OPCS4, UKB FID) | Definition |
|---|---|---|
| Chronic Kidney Disease Stage G3+ | ICD10 N180, N183 - N185, Y602, Y841, Z490, Z491, Z492, Z992; OPCS4 X40 to X42; FID 42027 | CKD stage 3 to stage 5, dialysis, algorithmically defined end-stage kidney disease, any eGFR < 60 mL/min/m2, uACR > 30 mg/mmol |
| Acute Kidney Injury | ICD10 N170 - N179 | Acute renal failure with tubular necrosis, acute renal failure with acute cortical necrosis, acute renal failure with medullary necrosis, other acute renal failure, acute renal failure (unspecified) |
| Ischemic Heart Disease | ICD10 I200, I21, I210, I211, I212, I213, I214, I219, I22, I220, I221, I228, I229, I23, I230, I231, I232, I233, I236, I238, I241, I252, I510, I513, I20, I201, I208, I209, I240, I251, Z951, Z955, I253, I254, I341, I25, I255, I256, I258, I259, I24, I248, I249 | Angina pectoris, unstable angina, myocardial infarction, coronary atherosclerosis, other acute and subacute forms of ischemic heart disease, other chronic ischemic heart disease (unspecified), aneurysm and dissection of heart |
| Cerebrovascular Disease | ICD10 G454, G463, G464, G465, G466, G467, G468, I600, I677, I679, I68, I680, I682, I688, I65, I650, I651, I652, I653, I658, I659, I631, I632, I672, I63, I632, I635, I639, I66, I660, I661, I662, I663, I664, I668, I669, I676, I630, I633, I634, I635, I636, I638, I64, I67, I678, G450, G451, G452, G458, G459, G460, G461, G462, I675, I671, I69, I690, I691, I692, I693, I694, I698 | Transient cerebral ischemia, cerebrovascular disease, occlusion of cerebral arteries, cerebral artery occlusion with cerebral infarction, occlusion and stenosis of precerebral arteries, cerebral ischemia, cerebral aneurysm, cerebral atherosclerosis, moyamoya disease, late effects of cerebrovascular disease |
| Diabetes Mellitus | ICD10 E10, E100, E106, E107, E108, E109, E101, E102, E103, E104, E11, E110, E116, E117, E118, E119, E13, E135, E136, E137, E138, E139, E149, E111, E131, E112, E103, E113, E123, E104, E114, E134, G590, Z964, R81, R824, G632, E103, E133, H360, R730, R739 | Type 1 diabetes, type 1 diabetes with ketoacidosis, type 1 diabetes with renal manifestations, type 2 diabetes with ophthalmic manifestations, diabetic retinopathy, type 1 diabetes with neurological manifestations, type 2 diabetes with ketoacidosis, type 2 diabetes with renal manifestations, type 2 diabetes with ophthalmic manifestations, type 2 diabetes with neurological manifestations, type 2 diabetes, polyneuropathy in diabetes, impaired fasting glucose, other abnormal glucose, glycosuria or acetonuria, insulin pump user |
| Hypertension | ICD10 I10, I13, I130, I131, I132, I139, I11, I110, I119, I12, I120, I129, I15, I150, I151, I152, I158, I159, I674 | Essential hypertension, hypertensive heart disease, hypertensive chronic kidney disease, hypertensive heart and/or renal disease, other hypertensive complications, |

**Table S3:** Genetic variants associated with TKV above genome-wide significance.[8]

See supplemental Excel file.

**Table S4:** Summary of GWAS studies used in two-sample MR analyses.

See supplemental Excel file.

**Table S5:** Baseline demographic characteristics

| **Phenotype[a, b]** | **Quantity** |
|---|---|
| TKV (mL) | 266.44 (74.5) |
| hTKV (mL/m) | 157.81 (37.55) |
| BSA TKV (mL/$m^2$) | 144.5 (29.15) |
| Age (y) | 64 (12) |
| Height (m) | 1.69 (0.14) |
| Weight (kg) | 74.9 (20.2) |
| BMI (kg/$m^2$) | 25.91 (5.32) |
| BSA ($m^2$) | 1.86 (0.3) |
| SBP (mmHg) | 139 (26) |
| eGFR Creatinine-Cystatin C (mL/min/1.73$m^2$) | 98.67 (17.62) |
| eGFR Creatinine (mL/min/1.73$m^2$) | 98.03 (15.82) |
| uACR (mg/mmol) | 1.07 (1.11) |
| HbA1C (%) | 5.32 (0.44) |
| Sex | |
| Female | 17835 (51.55) |
| Male | 16760 (48.45) |
| CKD Stage G3+ | 1805 (5.22) |
| Acute Kidney Injury | 499 (1.44) |
| Cerebrovascular Disease | 842 (2.43) |
| Ischemic Heart Disease | 2591 (7.49) |
| Type 2 Diabetes | 1414 (4.09) |
| Hypertension | 7131 (20.61) |
| Smoking | |
| Never | 21590 (62.41) |
| Previous | 11712 (33.85) |
| Current | 1180 (3.41) |
| Alcohol | |
| Never | 2084 (6.02) |
| <1 time/month | 3359 (9.71) |
| 1 to 3 times/month | 3962 (11.45) |
| 1 to 2 times/week | 9250 (26.74) |

| | |
|---|---|
| 3 to 4 times/week | 9998 (28.9) |
| Daily or almost daily | 5931 (17.14) |

a. Disease diagnoses include both incident and prevalent cases.
b. Sex, disease diagnoses, smoking, and alcohol consumption frequency are expressed as number of individuals and percentage of cohort. All other variables are expressed as median and interquartile range.

**Table S6:** Discrimination, calibration, and likelihood ratio test assessing CKD Stage G3+ risk prediction performance of eGFR and uACR compared to model incorporating TKV measurements.

| | C-Statistic | 95% CI | DeLong P[a] | Bias Corrected C-Statistic | Calibration P[a] | Bias Corrected Calibration P[a] | Likelihood Ratio Test P[a] |
|---|---|---|---|---|---|---|---|
| **eGFR + uACR** | 0.862 | 0.839 to 0.885 | Reference | 0.861 | 0.38 | 0.37 | Reference |
| **+TKV Thresholds** | 0.866 | 0.843 to 0.890 | 0.108 | 0.865 | 0.38 | 0.35 | 3.69E-05 |
| **+hTKV Thresholds** | 0.868 | 0.845 to 0.891 | 0.043 | 0.866 | 0.56 | 0.57 | 7.16E-06 |
| **+BSA TKV Thresholds** | 0.871 | 0.848 to 0.893 | 0.017 | 0.869 | 0.59 | 0.61 | 9.02E-08 |
| **+TKV** | 0.863 | 0.839 to 0.886 | 0.849 | 0.861 | 0.092 | 0.071 | 1.10E-02 |
| **+hTKV** | 0.863 | 0.840 to 0.887 | 0.588 | 0.862 | 0.084 | 0.087 | 3.24E-02 |
| **+BSA TKV** | 0.867 | 0.844 to 0.890 | 0.123 | 0.866 | 0.31 | 0.3 | 6.83E-06 |

a. Significance threshold corrected for multiple hypotheses testing = 0.05/3 or 0.017

**Table S7:** Improvement in category free risk reclassification when TKV, htTKV, or BSA-TKV were added to a baseline model of eGFR and uACR.

| Category Free Net Reclassification Index | %[a] | 95% CI | NARI[b] | P | Bias Corrected % | Bias Corrected NARI |
|---|---|---|---|---|---|---|
| **TKV** | | | | | | |
| **Event (n = 216)** | 15.74 | 2.57 to 28.91 | 1.12 | 0.019 | 15.67 | 1.11 |
| **Non-Event (n = 30,230)** | 1.02 | -0.11 to 2.14 | 10.08 | 0.077 | 1.05 | 10.39 |
| **Total (n = 30,446)** | 16.76 | 3.54 to 29.97 | 11.2 | 0.013 | 16.72 | 11.5 |
| **hTKV** | | | | | | |
| **Event (n = 216)** | 14.81 | 1.63 to 28.00 | 1.05 | 0.028 | 14.79 | 1.05 |
| **Non-Event (n = 30,230)** | 3.69 | 2.56 to 4.81 | 36.62 | <0.001 | 3.73 | 37.06 |
| **Total (n = 30,446)** | 18.5 | 5.27 to 31.74 | 37.67 | 0.006 | 18.52 | 38.11 |
| **BSA-TKV** | | | | | | |
| **Event (n = 216)** | 18.52 | 5.41 to 31.6 | 1.31 | 0.006 | 18.81 | 1.33 |
| **Non-Event (n = 30,230)** | 10.4 | 9.28 to 11.5 | 103.23 | <0.001 | 10.44 | 103.62 |
| **Total (n = 30,446)** | 28.92 | 15.76 to 42.1 | 104.54 | <0.001 | 29.24 | 104.95 |

a. Percentage calculated as difference between number of individuals with increased risk score (if in the event cohort) and those with decreased risk score divided by number of individuals in event, non-event, or total category.
b. Net absolute reclassification index was calculated as the product of reclassification percentage and event rate, multiplied by 1,000. Interpreted as absolute reclassification per 1,000 individuals.

**Table S8:** Inverse variance weighted, weighted median, and MR Egger regression results for each outcome associations using all SNPs.

See supplemental Excel file.

**Table S9:** Instrument strength, intercept test, and heterogeneity test results for each TKV-outcome association.

See supplemental Excel file.

**Table S10:** Inverse variance weighted, weighted median, and MR Egger results for each TKV-outcome association, excluding outlier SNPs identified by MR-PRESSO and BMI or BSA associated SNPs.

See supplemental Excel file.

**Table S11:** Nonlinear MR tests of nonlinearity and heterogeneity.[7]

See supplemental Excel file.

**Table S12:** Inverse variance weighted, weighted median, and MR Egger results for each genetically predicted exposure and TKV association.

See supplemental Excel file.

**Table S13:** Instrument strength, intercept test, and heterogeneity test results for each exposure-outcome association.

See supplemental Excel file.

**Table S14:** Inverse variance weighted, weighted median, and MR Egger results for each genetically predicted exposure and TKV association, excluding outlier variants identified by MR-PRESSO.

See supplemental Excel file.
